# Clinical significance of IgM and IgG test for diagnosis of highly suspected COVID-19 infection

**DOI:** 10.1101/2020.02.28.20029025

**Authors:** Xingwang Jia, Pengjun Zhang, Yaping Tian, Junli Wang, Huadong Zeng, Jun Wang, Jiao Liu, Zeyan Chen, Lijun Zhang, Haihong He, Kunlun He, Yajie Liu

## Abstract

Quick, simple and accurate diagnosis of suspected COVID-19 is very important for the screening and therapy of patients. Although several methods were performed in clinical practice, however, the IgM and IgG diagnostic value evaluation was little performed. 57 suspected COVID-19 infection patients were enrolled in our study. 24 patients with positive and 33 patients with negative nucleic acid test. The positive rate of COVID-19 nucleic acid was 42.10%. The positive detection rate of combination of IgM and IgG for patients with COVID-19 negative and positive nucleic acid test was 72.73% and 87.50%. The results were significantly higher than the nucleic acid or IgM, IgG single detection. hsCRP in the COVID-19 nucleic acid negative group showed significantly higher than the positive groups (*P*=0.0298). AST in the COVID-19 IgM negative group showed significantly lower than the positive groups (*P*=0.0365). We provided a quick, simple, accurate aided detection method for the suspected patients and on-site screening in close contact with the population.

## Introduction

COVID-19 which was discovered in Wuhan due to pneumonia virus cases in 2019[1], and was named by the World Health Organization on January 12, 2020. Coronaviruses are a large family of viruses that are known to cause colds and more serious diseases[2]. COVID-19 is a novel coronavirus strain that has never been found in humans before. Common signs of a person infected with COVID-19 include respiratory symptoms, fever, cough, shortness of breath, and dyspnea. In more severe cases, infection can cause pneumonia, severe acute respiratory syndrome, kidney failure, and even death. There is currently no specific treatment for diseases caused by COVID-19[3]. However, many symptoms can be managed, so they need to be treated according to the clinical situation of the patient. The main routes of transmission of COVID-19 are respiratory droplets and contact transmission. Aerosol and fecal-oral routes of transmission need to be further clarified. Epidemiological investigations have shown that cases can be traced to close contact with confirmed cases[4, 5].

According to the sixth edition of the diagnostic criteria, the COVID-19 cases are divided into two categories: “suspected cases” and “confirmed cases”. As of 24:00 on February 25, a total of 77,789 confirmed cases have been reported in China, 27,836 cases have been cured, 2,666 death cases. COVID-19 is sudden and public events in the worldwide[6]. Timely and accurate diagnosis of it is very important for the detection and therapy of patients. However, in the clinical practice, the detection standard varied partly with rapidly growing awareness of COVID-19. Nucleic acid detection, chest CT, epidemiological history and clinical manifestations were recognized as diagnostic basis[7, 8]. However, the nucleic acid detection had the limitation of operators, time consuming, easy pollution. CT results often can be changed at severely infected patients, and the specificity was also limited. IgM/IgG antibody detection method has the advantages of simple, easy, and high sensitivity.

In our study, by evaluating the clinical significance of IgM and IgG for the highly suspected COVID-19 infection patients, we aimed to provide a quick, simple, accurate diagnostic method for the detection of suspected patients.

## Materials and Methods

### Patients

This is a retrospective study which was approved by the Ethics Committee of Shenzhen Hospital, Southern Medical University (NYSZYYEC20200009). The data were anonymous, so the requirement for informed consent was therefore waived. Total 57 suspected COVID-19 infection patients were enrolled in our study according National Health Commission of the people’s Republic of China. Diagnostic and treatment protocol for COVID-19 (trial Sixth Edition). Definition of suspected cases of COVID-19: first, at least one of the following clear epidemiological history: (1) the patient has a history of travel or resident in Wuhan or surrounding area, or communities with COVID-19 patients within 14 days before onset; (2) has a contact history with people infected with COVID-19 (positive nucleic acid test) within 14 days before onset; (3) has a contact history with patients from Wuhan and surrounding areas, or has a contact history with patients who has fever or respiratory symptoms from communities with COVID-19; (4) Cluster onset. And at least have the following two clinical manifestations: (1) fever and (or) respiratory symptoms; (2) conforming to the imaging features; (3) white blood cells are normal or reduced in early stage of disease, and lymphocyte count is reduced. Second, if there is no clear epidemiological history, it meets the above three clinical manifestations.

### Laboratory Examination

Blood routine and hs-CRP were detected by Mindray CAL8000 automatic blood analyzer (Mindray Company, Shenzhen). The hs-CRP(Lot : 2019111901) detection method was immunoturbidimetric method. The AST (Lot: 252968) and ALT (Lot: 021837) used the VITROS V5600 automatic biochemical immunoassay analyzer (Ortho, Rochester, NY). The methods were dry chemical methods. D-Dimer (Lot:255469) was detected by STAGO-R MAX automatic hemagglutination analyzer (Diagnostic Stago, Gennevilliers), and the detection method was immunoturbidimetry. Primary screening of pharyngeal swab nucleic acid amplification was performed by two kits of 6 companies (DAAN, Sansure Biotech, BGI, ShangHai ZJ Biotech, Geneodx, Biogerm) in more than 20 hospitals of ShenZhen. COVID IgM/IgG antibodies kit which have sent to BIMT for product verification were detected on Time-Resolved Immunofluorescence Analyzer by Fluorescence immunochromatographic assay method (Beijing Diagreat Biotechnologies Co., Ltd,Lot: 20200214). The procedure of nucleic acid, IgM and IgG detection was strictly performed according to the instruction of manufacturer’s manual. 242 healthy people without related diseases were tested, and the values were measured in ascending order. Among them, 95% of the values were negative. Then we defined the cutoff of IgM and IgG are 0.88 and 1.02. The result were expressed as fluorescence intensity (Flu).

### Data analysis

Statistical analyses were performed with the Statistical Analysis System software SPSS 19.0, and data are presented as Median (25% percentile, 75% percentile). With nonparametric test and two-sided χ2 test, we compared the differences between the two groups, and the *P*-value <0.05, which will be considered statistically significant.

## Results

### Clinical characteristics and COVID-19 nucleic acid tests

According to the diagnostic standard of suspected COVID-19 infection, 57 patients were enrolled in our study. All the 57 patients underwent 3 times nucleic acid tests, and every time the results of nucleic acid tests were confirmed by two COVID-19 nucleic acid tests kits. Of all the 57 patients, 24 patients had a positive nucleic acid test and 33 patients had a negative nucleic acid test for the first time, and all the 57 patients had a negative nucleic acid test for the second and third time. The positive rate of COVID-19 nucleic acid in the 57 suspected COVID-19 infection was 42.10%. From the time of the first exposure to COVID-19 infection to the nucleic acid test, the time ranged from 1 day to 34 days. 1 patient had negative nucleic acid result after 34 days exposure, however, the IgM detection result was positive. The results were partly different from the current cognition “The median incubation period for COVID-19 is 3 days, with a minimum of 0 days and a maximum of 24 days”.

### IgM and IgG single detection for COVID-19

According to the nucleic acid test results, we performed IgM and IgG detection by the Diagreat company. As shown in Figure 1A, among the 33 patients with COVID-19 nucleic acid negative results, the IgM fluorescence intensity (Flu) of 20 patients was more than 0.88, the positive rate was 60.61%. As shown in Figure 1B, the IgG Flu of 15 patients was more than 1.02, the positive rate was 45.45%. As shown in Figure 2A, among the 24 patients with COVID-19 nucleic acid positive results, the IgM Flu of 19 patients was more than 0.88, the positive rate was 79.17%. As shown in Figure 2B, the IgG Flu of 16 patients was more than 1.02, the positive rate was 66.67%.

**Figure 1.**
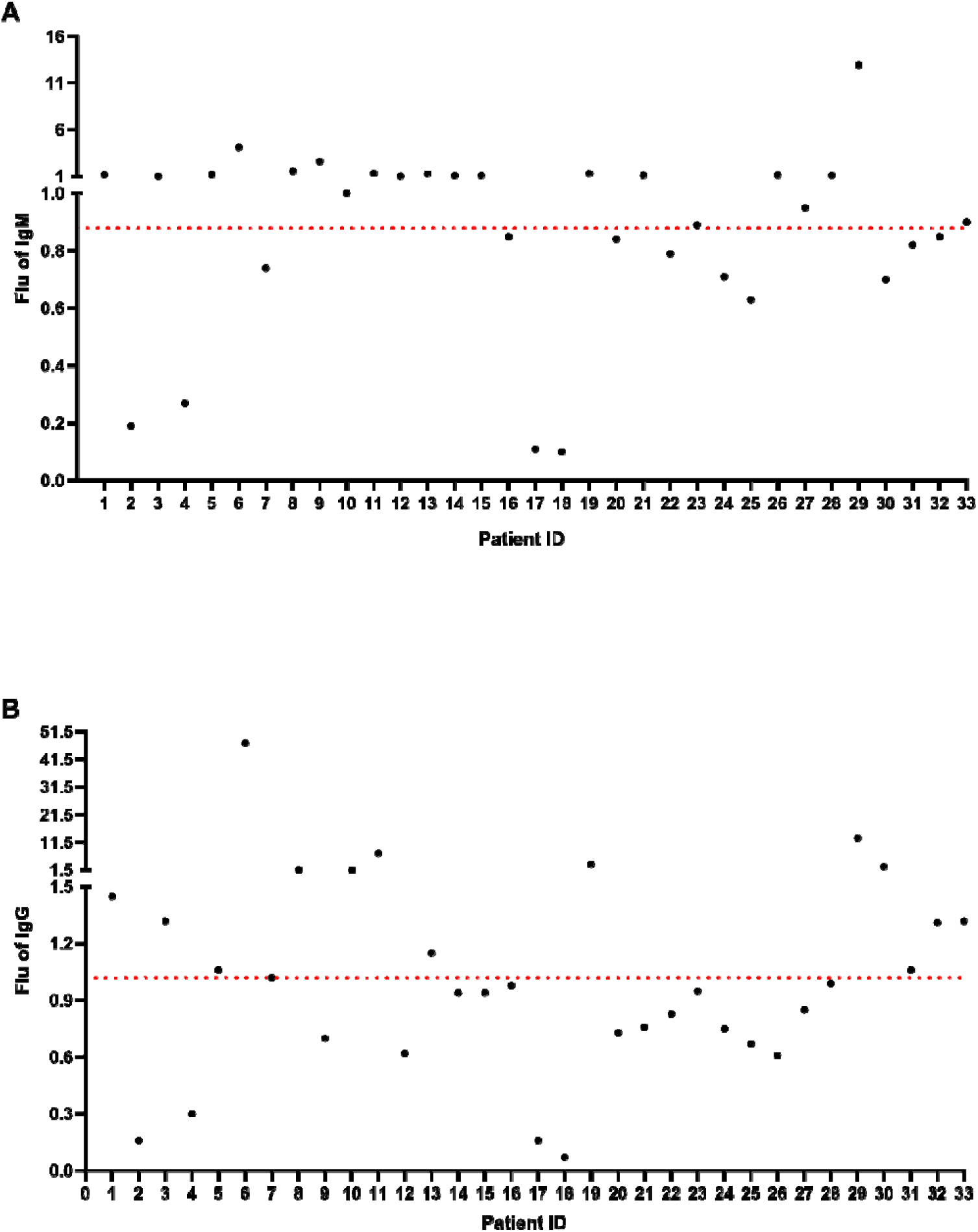
IgM and IgG detection among the 33 patients with COVID-19 nucleic acid negative results. A:IgM fluorescence intensity (Flu) of 20 patients was more than 0.88. B, IgG Flu of 15 patients was more than 1.02.

**Figure 2.**
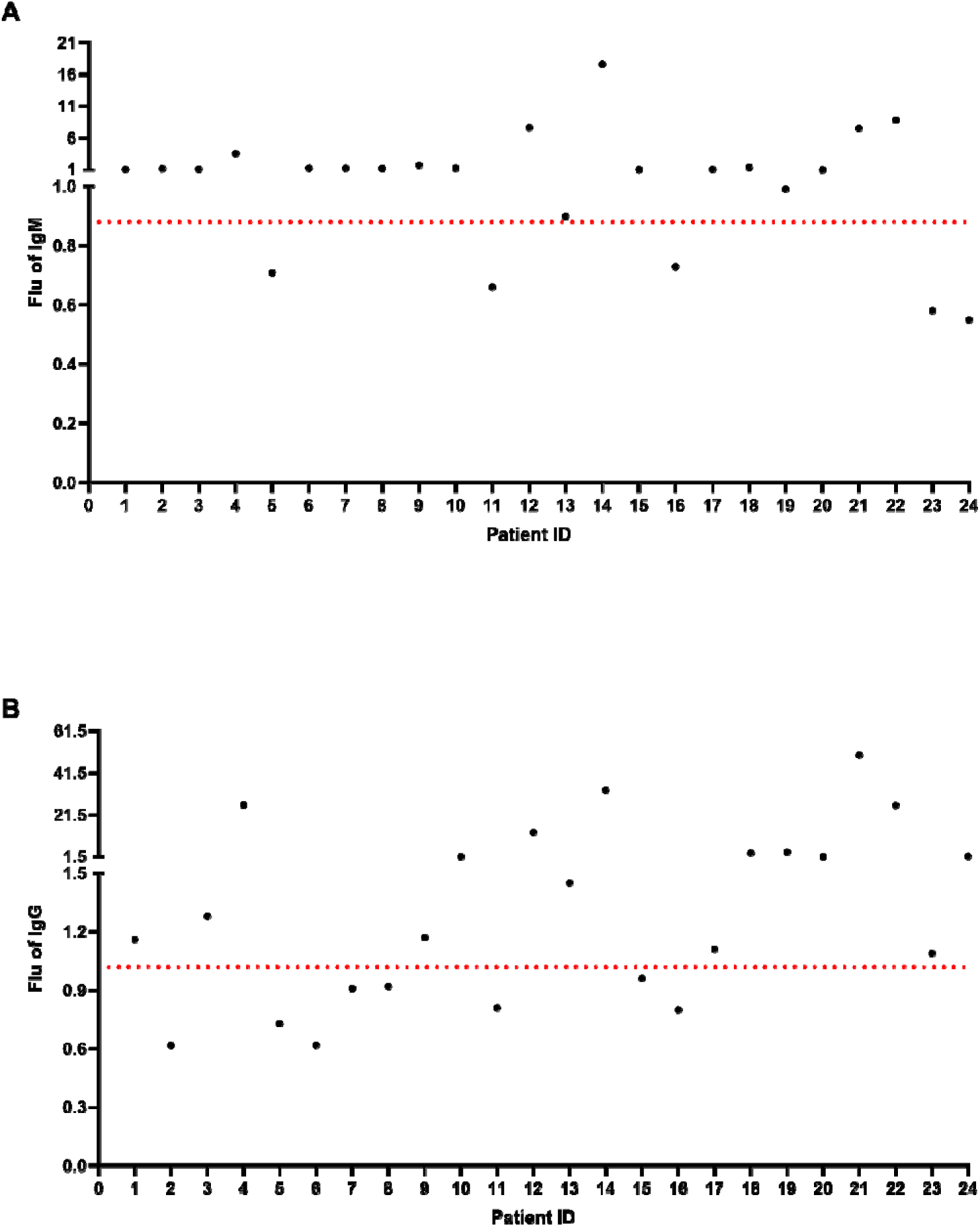
IgM and IgG detection among the 24 patients with COVID-19 nucleic acid positive results, A:IgM fluorescence intensity (Flu) of 19 patients was more than 0.88. B, IgG Flu of 16 patients was more than 1.02.

### Combination of IgM and IgG detection for COVID-19

As shown in Figure 3A, combination of IgM and IgG detection for COVID-19, during the 33 patients had a negative nucleic acid test, the percentage rate of IgM(+)IgG(+), IgM(-)IgG(+), IgM(+)IgG(-), IgM(-)IgG(-) were 36.37%, 12.12%, 24.24%, and 27.27%, separately. The positive diagnostic rate of combination of IgM and IgG detection for 33 patients with COVID-19 negative nucleic acid test was 72.73%.

**Figure 3.**
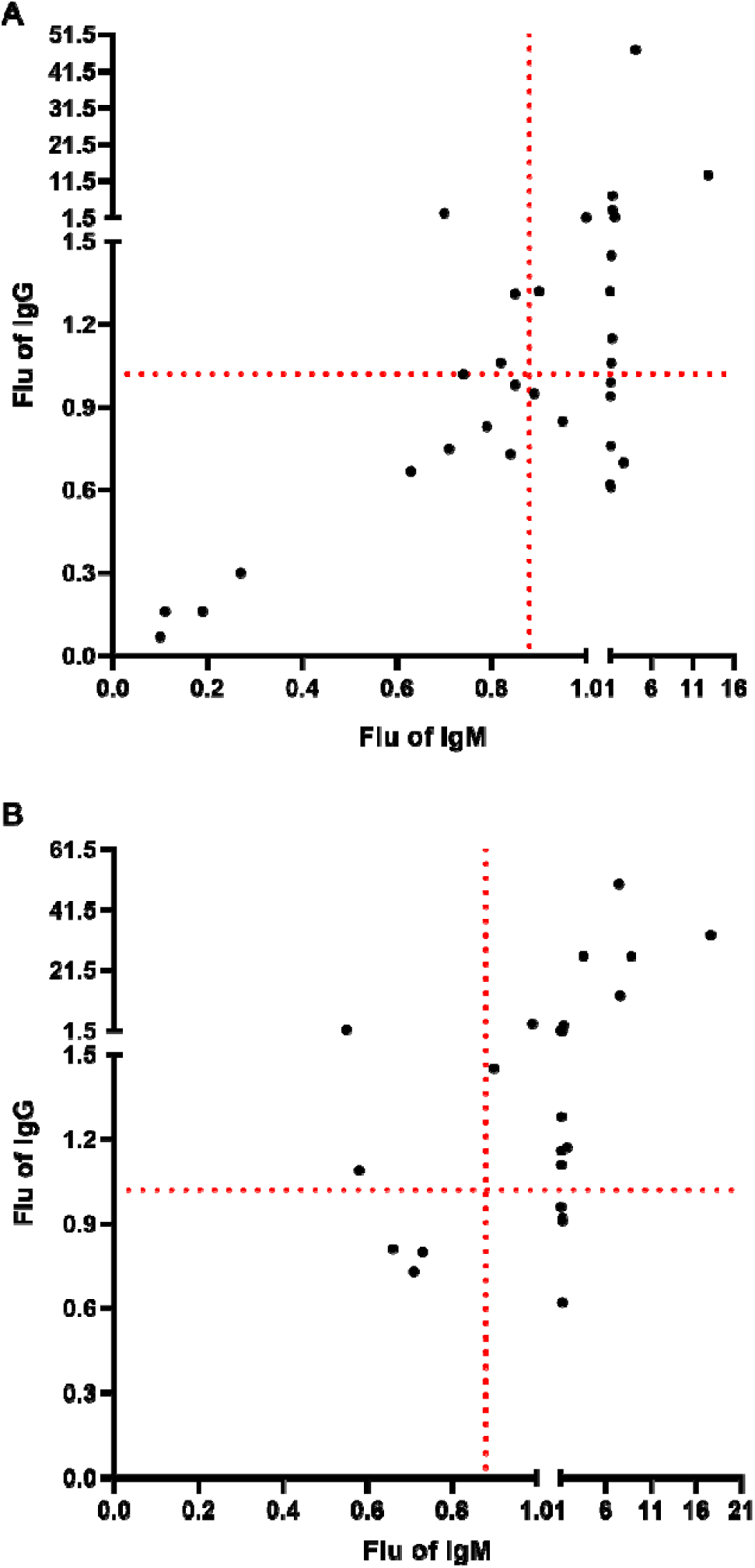
Combination of IgM and IgG detection for COVID-19. A: The positive diagnostic rate of combination of IgM and IgG detection for 33 patients with COVID-19 negative nucleic acid test was 72.73%. B: The positive diagnostic rate of combination of IgM and IgG detection for 24 patients with COVID-19 negative nucleic acid test was 87.50%.

Compared with the negative nucleic acid test, IgM and IgG single detection, the combination of IgM and IgG showed significantly increased (*P*<0.01). As shown in Figure 3B, combination of IgM and IgG detection for COVID-19, during the 24 patients had a positive nucleic acid test, the percentage rate of IgM(+)IgG(+), IgM(-)IgG(+), IgM(+)IgG(-), IgM(-)IgG(-) were 62.50%, 8.33%, 16.67%, and 12.50%, separately. The positive diagnostic rate of combination of IgM and IgG detection for 24 patients with COVID-19 negative nucleic acid test was 87.50%. Compared with the nucleic acid positive test, IgM and IgG single detection, the combination of IgM and IgG also showed significantly increased (*P*<0.01).

### Blood indicators for COVID-19 nucleic acid, IgM and IgG test

Nine blood indicators (hsCRP, WBC, LY, LY%, NEUT, NEUT%, PLT, ALT and AST) were analyzed for the negative and positive group of COVID-19 nucleic acid, IgM and IgG test. As shown in Figure 3, the concentration of hsCRP in the COVID-19 nucleic acid negative and positive groups were 1.10 (0.36, 2.43) and 0.51(0.20, 0.77), separately. It showed significantly difference between the two groups (*P*=0.0298). The other eight indicators (WBC, LY, LY%, NEUT, NEUT%, PLT, ALT and AST) showed not significantly difference (*P*>0.05). As shown in Figure 4, the concentration of AST in the COVID-19 IgM negative and positive groups were 21.00 (17.00, 26.75) and 23.00(21.00, 32.00), separately. It showed significantly difference between the two groups (*P*=0.0365). The other eight indicators (hsCRP, WBC, LY, LY%, NEUT, NEUT%, PLT, and ALT) showed not significantly difference (*P*>0.05). As shown in Figure 5, all the nine indicators showed not significantly difference (*P*>0.05) between the COVID-19 IgG negative and positive groups.

**Figure 4:**
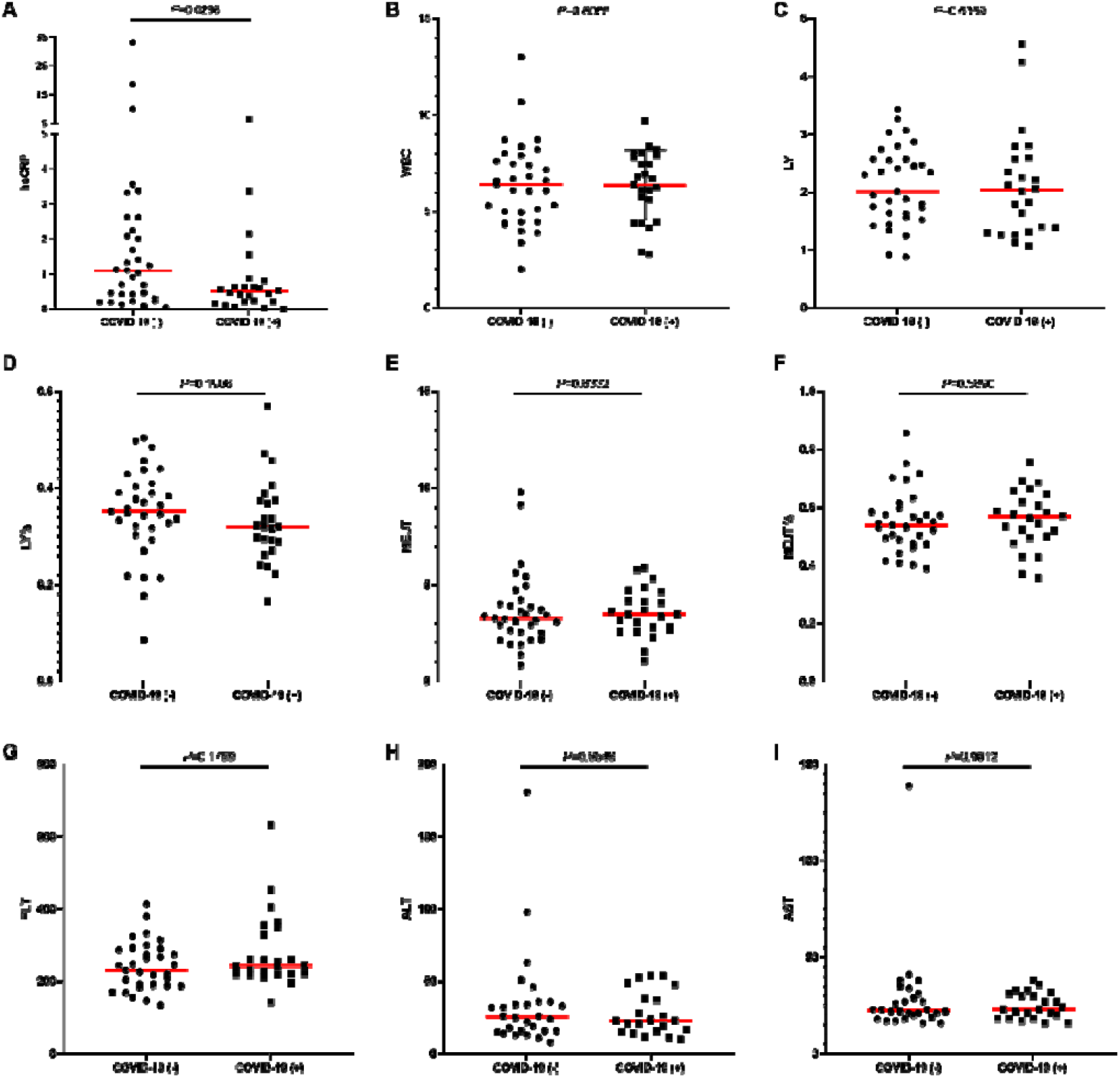
Nine blood indicators were analyzed for the negative and positive group of COVID-19 nucleic acid test. A: hsCRP, B:WBC, C:LY, D: LY%, E: NEUT, F:NEUT%, G: PLT, H: ALT, I:AST.

**Figure 5:**
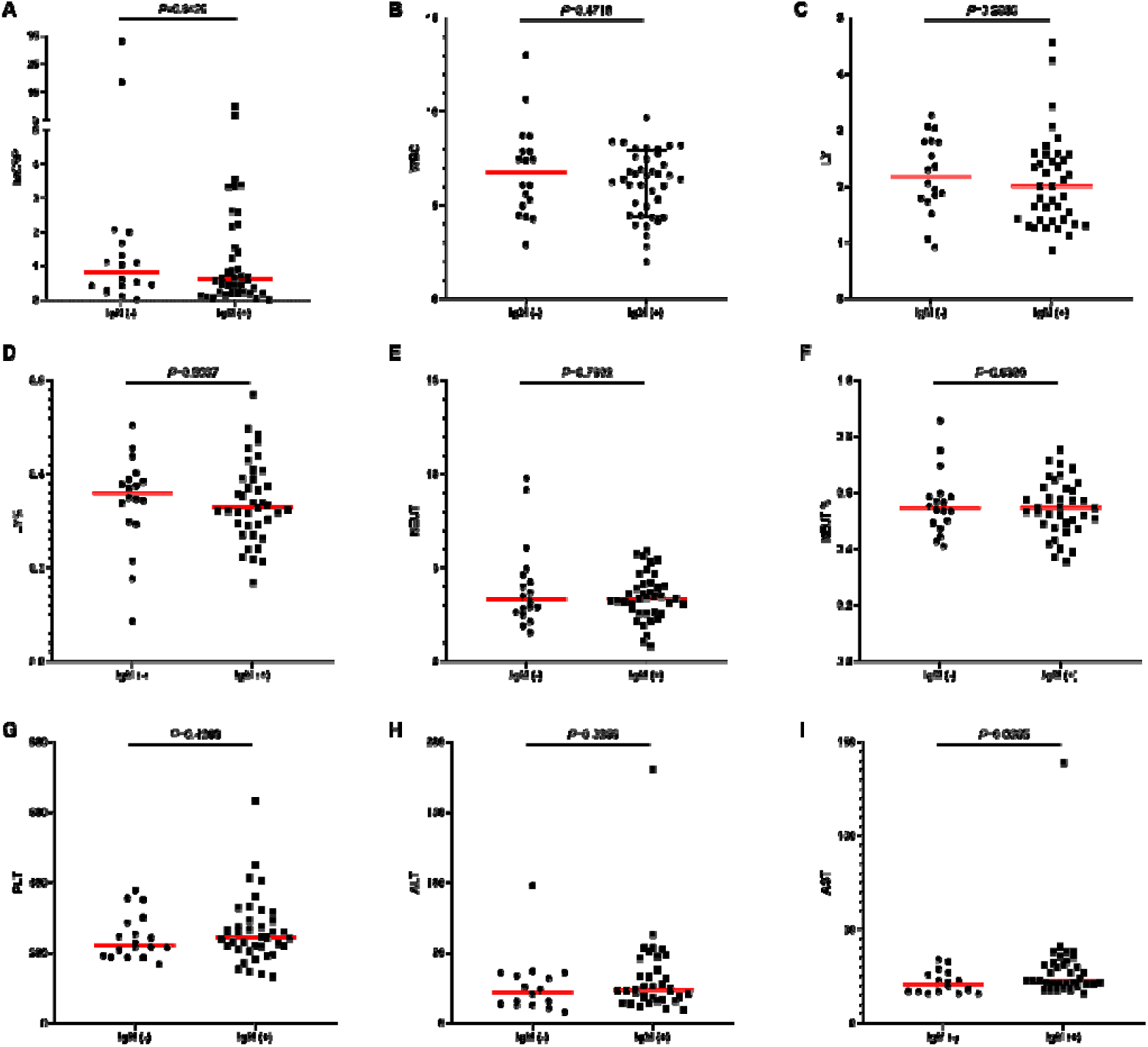
Nine blood indicators were analyzed for the negative and positive group of IgM test. A: hsCRP, B:WBC, C:LY, D: LY%, E: NEUT, F:NEUT%, G: PLT, H: ALT, I:AST.

## Discussion

In our study, the positive rate of COVID-19 nucleic acid in the 57 suspected COVID-19 infection was 42.10%. The results of nucleic acid results may be false negative and also false positive. First, because of the throat swab samples were collected in our study. Studies demonstrated that the sample location was a very important factor for the nucleic acid detection. Researchers analyzed a total of 72 nasal swabs and 72 throat swabs to obtain 9 consecutive samples from each patient. They found that a higher viral load was detected shortly after the onset of symptoms, with viral load in the nose being higher than in the throat[9]. Liu YB collected each 100 nasal and throat swab specimens. of which 89 specimens were positive for the nasal swab specimens, and 54 specimens were positive for the throat swab specimens. The positive rate in nasal swab samples was significantly higher than that in throat swab samples. Second, in the clinical practice, when collecting throat swab specimens, medical staff need wear protective clothing. Different medical staff operated the sample collecting. The procedure may also affect the nucleic acid detection results. As shown in Figure 7A, the nucleic acid detection result of Patient ID 55 was negative, but the IgM and IgG results were positive. In the lower of both lungs, there were large fuzzy shadows and GGO, some slightly fan-shaped distribution. Third, because of the laboratory pollution, the positive results may be also false positive. As shown in Figure 7B, the nucleic acid detection result of Patient ID 19 was positive, but the IgM and IgG were negative results, according to the CT results, no obvious lesion was found in both lungs. Study analyzed 126 German citizens left Wuhan who had to pass screening for clinical signs of infection. 2 passengers nucleic acid test were positive after quarantine for 14 days, but the two patients did not develop symptoms. The researchers reconfirmed the results by other methods. The result indicate that people with no fever, no symptoms, or only mild symptoms of infection may ignore their potential infectivity[10]. Laboratory examination of the COVID-19 nucleic acid positive group of 31 patients, negative group of 23 patients are mainly characterized by reduced lymphocyte counts, increased C-reactive protein. Except for dyspnea, there was no significantly different in the clinical characteristics of the covid-19 nucleic acid negative and positive group[11]. Studies demonstrated that clinical features of clinical diagnosis of COVID-19 nucleic acid positive and negative patients are similar, faster and more accurately methods were urgently needed for the diagnosis of COVID-19. The positive diagnosis rate of combination of IgM and IgG detection for patients with COVID-19 negative and positive nucleic acid test was 72.73% and 87.50%. Although several IgM and IgG detection kit promotion were reported by news, however, little study was reported. One study enrolled 6 cases of COVID-19 Gansu Province were analyzed by COVID-19 specific IgM antibody detection kit. 5 cases were positive and 1 case was negative[12].

**Figure 6:**
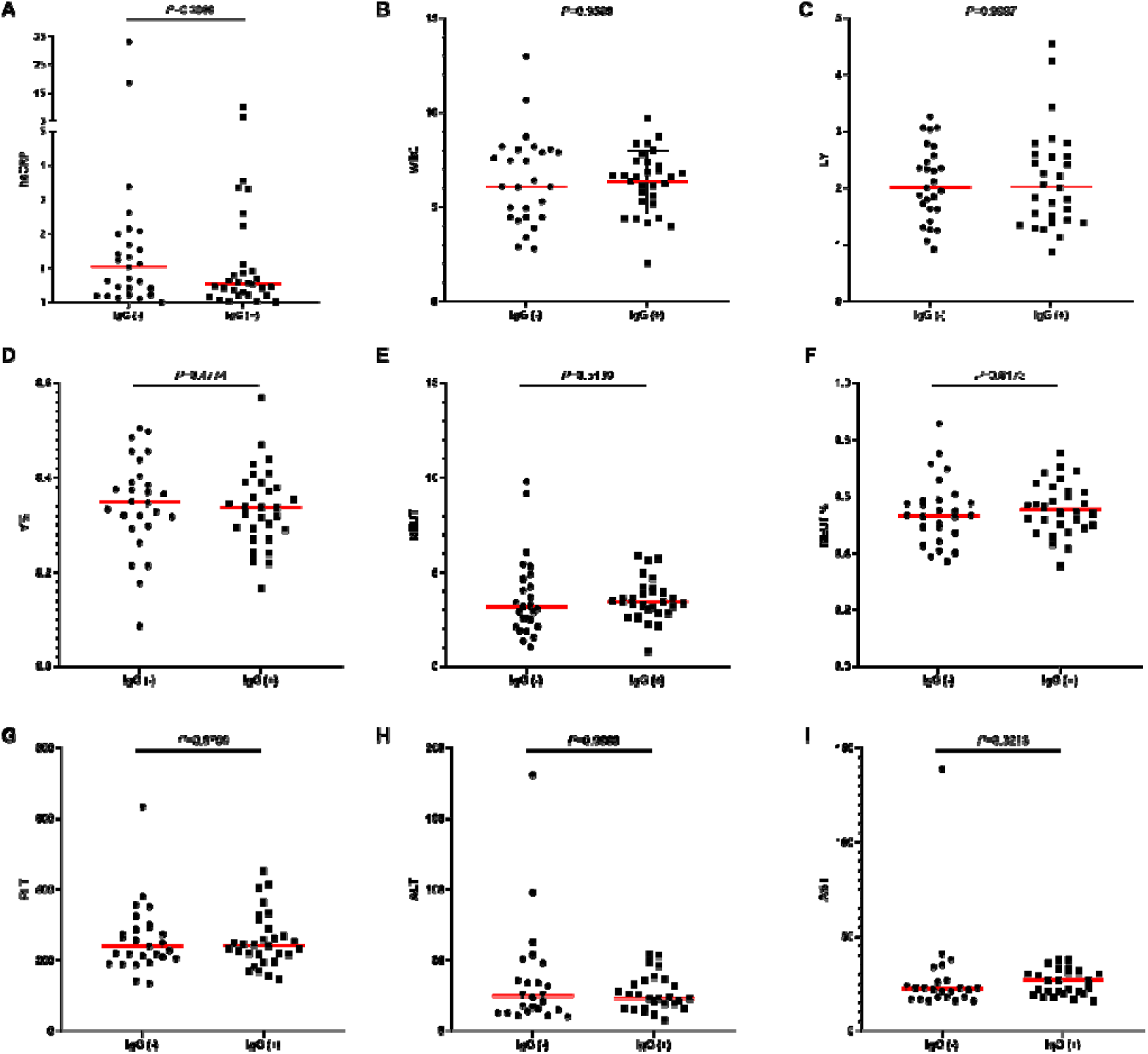
Nine blood indicators were analyzed for the negative and positive group of IgG test. A: hsCRP, B:WBC, C:LY, D: LY%, E: NEUT, F:NEUT%, G: PLT, H: ALT, I:AST.

**Figure 7.**
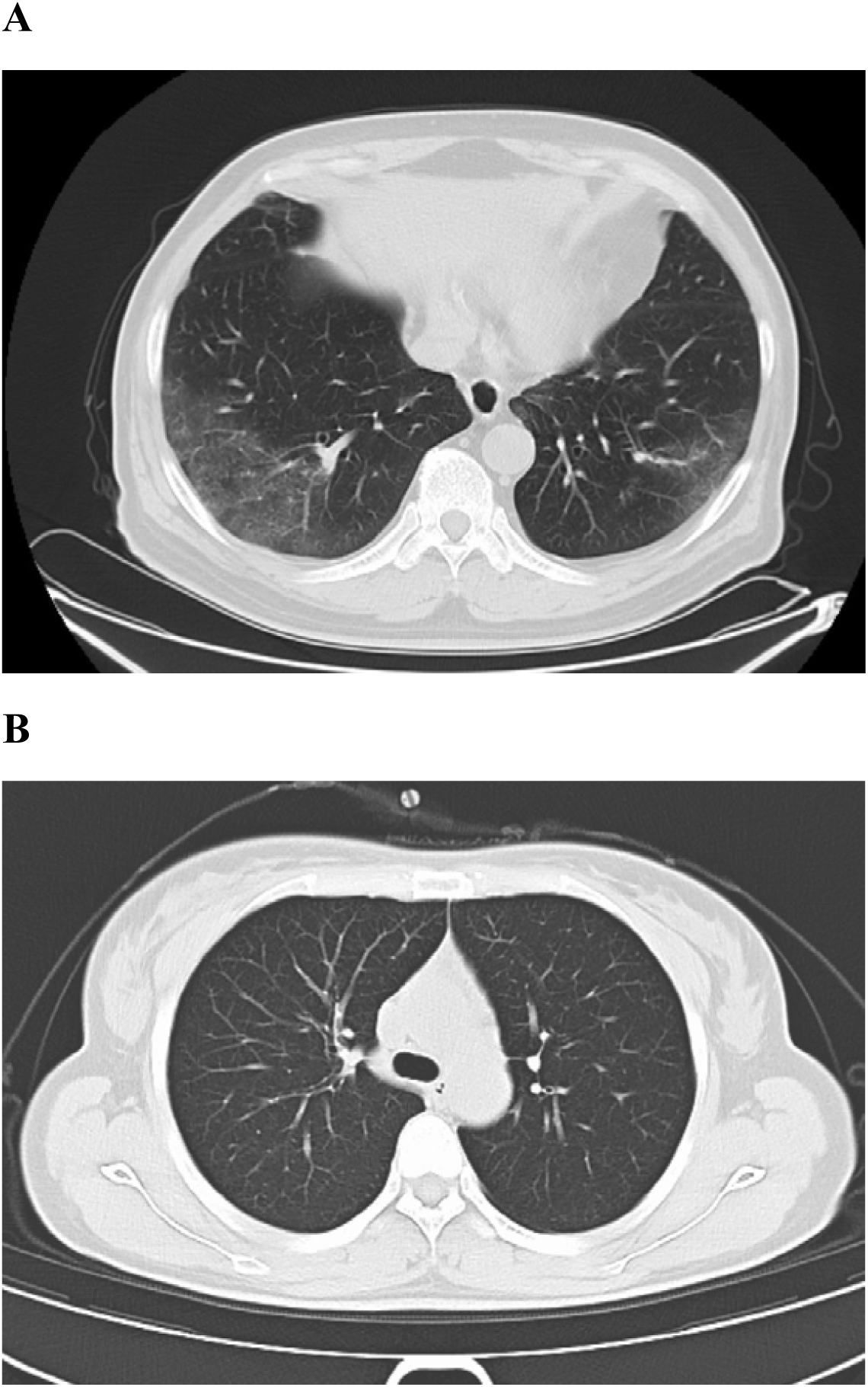
Patient case CT scan. A: Patient ID 55, the nucleic acid detection result was negative, but the IgM and IgG results were positive. In the lower of both lungs, there were large fuzzy shadows and GGO, some slightly fan-shaped distribution. B: Patient ID 19, the nucleic acid detection result was positive, but the IgM and IgG results were negative, no obvious lesion was found in both lungs.

Studies analyzed coagulation indicators and outcomes of consecutive 183 patients with confirmed COVID-19, found significantly elevated D-dimer and fibrin degradation levels are closely related with the deaths[13]. Study analyzed the clinical symptoms of 20 medical workers infected with COVID-19, Compared with normal patients, white blood cell count and liver enzyme showed significantly increased in severe patients, the lymphocyte count in peripheral blood were significantly decreased[14]. In our study, the concentration of hsCRP in the COVID-19 nucleic acid negative and positive groups were 1.10 (0.36, 2.43) and 0.51(0.20, 0.77), separately. It showed significantly difference between the two groups (P=0.0298). C-reactive protein is an acute phase protein synthesized by liver cells when the body is exposed to inflammatory stimuli such as microbial invasion or tissue damage. The detection of CRP is widely used in clinical applications, including the diagnosis and differential diagnosis of acute infectious diseases, the monitoring of postoperative infections, the observation of the efficacy of antibiotics, the detection of disease course and prognosis. In clinical practice, some patients with COVID-19 would suddenly worsen in the later stage and soon enter a state of multiple organ failure. It may be related to the sudden initiation of an inflammatory storm in critically ill patients. The inflammatory storm not only causes lung injury, but also causes multiple organs such as the liver, heart muscle, and kidney damage. Therefore, “inflammatory storm” is also one of the important reasons for liver injury in COVID-19 patients. In our study, the concentration of AST in the COVID-19 IgM negative and positive groups were 21.00 (17.00, 26.75) and 23.00(21.00, 32.00), separately. It showed significantly difference between the two groups (*P*=0.0365). Studies demonstrated that the liver function indicators of patients with COVID-19 intensive care unit were significantly higher than those in non-intensive care units[15, 16]. In another 1099 patients from multiple centers study, it found that ALT and AST were also mainly elevated in severe patients[17]. Studies revealed that COVID-19 enters cells mainly through angiotensin converting enzyme 2. Bile duct epithelial cells specifically express ACE2, which is 20 times higher than hepatocytes, suggesting that 2019-nCoV infection may cause bile duct epithelial cell damage[18]. The reason of liver function abnormalities is more mainly due to drugs, systemic inflammation, and multiple organ dysfunction Secondary liver damage, not liver damage caused by the virus itself[19].

There are still some limitations in our study. First, the relatively small sample size, difference of IgM and IgG antigen binding site, difference of COVID-19 nucleic acid design, it may result in the bias of results. Second, because of the different time that from a patient who were firstly exposed to the virus to the detection, the detection positive rate of IgM and IgG may be affected. Earlier and different times should be performed to validate the detection value of IgM and IgG. Third, the detection value of IgM and IgG should be followed up in the future study.

In summary, compared with the nucleic acid detection, the IgM and IgG may provide a quick, simple and accurate aided detection method for suspected COVID-19 patients. We should combine the nucleic acid, IgM, IgG, CT scan and clinical characteristics results together for the diagnosis of COVID-19.

## Data Availability

The data used to support the findings of this study are available from the corresponding author upon request.

## Author Contribution

Xingwang Jia, Yaping Tian, Jun Wang, Kunlun He, and Yajie Liu contributed to the study design. Junli Wang and Huadong Zeng contributed to data collection. Jiao Liu and Haihong He contributed to the collection of clinical specimens. Xingwang Jia, Zeyan Chen and Lijun Zhang contributed to experiments and data collection. Pengjun Zhang contributed to the data analysis. Xingwang Jia, Pengjun Zhang and Yaping Tian contributed to the manuscript preparation.

## Acknowledgments

We thank Professor Fabao Gao of West China Hospital of Sichuan University and Dr. Linfen Zhao of Nanjing University of Chinese Medicine Affiliated Wujin Hospital of Traditional Chinese Medicine help us revise the sentences in English description of CT imaging.

